# Tocilizumab in hospitalized patients with COVID-19: A meta analysis of randomized controlled trials

**DOI:** 10.1101/2021.03.23.21254054

**Authors:** Vijairam Selvaraj, Mohammad Saud Khan, Kwame Dapaah-Afriyie, Arkadiy Finn, Chirag Bavishi, Amos Lal, Eleftarios Mylonakis

## Abstract

**Background:** To date, only dexamethasone has been shown to reduce mortality in COVID-19 patients. Tocilizumab has been recently added to the treatment guidelines for hospitalized COVID-19 patients, but data remains conflicting.

**Methods:** Electronic databases such as MEDLINE, EMBASE and Cochrane central were searched from March 1, 2020, until February 28th, 2021, for randomized controlled trials evaluating the efficacy of tocilizumab in hospitalized COVID-19 patients. The outcomes assessed were all-cause mortality at 28 days, mechanical ventilation, and time to discharge.

**Results:** Eight studies (with 6,311 patients) were included in the analysis. In total, 3,267 patients received tocilizumab, and 3,044 received standard care/placebo. Pooled analysis showed a significantly decreased risk of all-cause mortality at 28 days (RR 0.90, 95% CI 0.83-0.97, p=0.009) and progression to mechanical ventilation (RR 0.79, 95% CI 0.70-0.90, p=0.0002) in the tocilizumab arm compared to standard therapy or placebo. In addition, there was a trend towards improved median time to hospital discharge (RR 1.18, 95% CI 1.05-1.34, p=0.007).

**Conclusions:** Tocilizumab therapy improves outcomes of mortality and need for mechanical ventilation, in hospitalized patients with COVID-19 infection compared with standard therapy or placebo. Our findings suggest the efficacy of tocilizumab therapy in hospitalized COVID-19 patients and strengthen the concept that tocilizumab is a promising therapeutic intervention to improve mortality and morbidity in COVID-19 patients.

## INTRODUCTION

The coronavirus disease-19 (COVID-19) pandemic has resulted in hospitalization in many cases. In severe and critical cases of COVID-19, which occurs in 13.8% and 6.1% of the patient population, COVID-19 associated pneumonia can lead to acute respiratory distress syndrome (ARDS) and rapid deterioration, sometimes leading to invasive mechanical ventilation and death [1]. The clinical spectrum of COVID-19 continues to evolve along with the emergence of new severe acute respiratory syndrome-coronavirus-2 (SARS-CoV2) variants.

The pathophysiology of COVID-19 involves an initial viremic phase where patients mostly have mild constitutional symptoms, followed by a pulmonary and then hyperinflammatory phase where patients have shortness of breath and hypoxemia [2]. The median time from onset of symptoms to hospital admission is five to seven days and nine to ten days for acute respiratory distress syndrome (ARDS) and hypoxic respiratory failure [3,4]. The hyperinflammatory phase of COVID-19 is associated with elevated C-reactive protein (CRP) levels, ferritin, lactate dehydrogenase (LDH), and interleukin-6 (IL-6) and causes edema and inflammatory cell infiltration in the lungs [5,6]. IL-6 appears to play a significant role in endothelial dysfunction and the development of vascular permeability and has been associated with the vascular dysfunction seen in severe disease [7]. This dysregulated and excess immune response plays an important role in the disease course of COVID-19 [8,9]. Elevated levels of IL-6 have been associated with prolonged viral shedding, increased viremia, and progression to mechanical ventilation and death [10-13]. A meta-analysis of 6 studies revealed that mean IL-6 levels were 2.9-fold higher in patients with complicated COVID-19 than non-complicated disease [14].

Dexamethasone therapy has shown mortality benefit in patients of COVID-19 infection [15,16]. Nevertheless, in some severely ill patients, dexamethasone therapy alone might not be sufficient to quell the cytokine storm in the hyperinflammatory phase. Tocilizumab is a recombinant monoclonal antibody indicated for treating giant cell arteritis, rheumatoid arthritis, and life-threatening cytokine release syndrome induced by chimeric antigen receptor T-cells [17-19]. Its mode of action is by inhibiting IL-6 signaling by binding soluble IL-6R and membrane IL-6R [20]. A recent study also showed there might be genetic variants in the interleukin-6 inflammatory pathway that may be associated with life-threatening disease in COVID-19 patients [21]. It is reasonable to assume that early intervention with tocilizumab through IL-6 blockade could abrogate progression to hypoxemic respiratory failure and decreases the duration of supplemental oxygen use [22]. However, randomized controlled trials (RCTs) and systematic reviews evaluating the role of tocilizumab in COVID-19 patients have yielded disparate results 23-29].

We, therefore, conducted a meta-analysis of the RCTs to synthesize the current evidence on the efficacy of tocilizumab in hospitalized COVID-19 patients.

## METHODS

### Data Sources and Search Strategy

This study was conducted according to the Cochrane Collaboration and the Preferred Reporting Items for Systematic reviews and Meta-Analyses (PRISMA) statement [30]. We conducted a systematic search in MEDLINE, EMBASE, Cochrane Central and preprint databases to identify all relevant articles using the following search terms: (“SARS-CoV2” OR “COVID-19”) AND (“tocilizumab” OR “IL6” OR “Anti-IL6”). Results were limited to humans and the English language. Databases were searched from March 1, 2020, to February 28, 2021, to identify all relevant RCTs including preprint, non-peer reviewed studies as well. Review articles, case reports, observational studies, opinion articles, letters, abstracts, conferences, brief reports, and non-English publications were excluded. The reference lists of the identified articles were also perused to find additional pertinent studies. All results were imported into EndNote x8.2 (Clarivate Analytics) and duplicate results were identified and removed.

### Study Selection and Eligibility

Two independent reviewers (V.S. and M.S.K.) screened the retrieved papers based on the title and abstract. If the paper contained relevant data, the full paper was retrieved if it was not clear from the title and abstract. Any disagreements between the two reviewers were discussed with a third reviewer (K.D.A.) and resolved by consensus. A study was considered eligible for inclusion in the analysis if it was 1) randomized controlled trial 2) reported outcomes of interest in hospitalized COVID-19 patients with tocilizumab therapy compared to standard treatment or placebo. The outcomes assessed were all-cause mortality at 28 days, progression to mechanical ventilation, and the median time to hospital discharge. If more than one study reported data from the same population, then the largest study was included.

### Data extraction and quality assessment

From the included studies, two reviewers (V.S. and M.S.K.) independently extracted data. Extracted data included 1) study characteristics - design, site of study, dates of study, and type of randomization 2) details of the study population and the interventions utilized, including demographics of participants in both intervention and control arms, presence of comorbidities, concomitant treatment, CRP levels 3) primary outcome and follow up.

### Risk of Bias Assessment

Two authors (V.S. and M.S.K.) reviewed each selected trial for quality assessment, including the risk of bias using the Cochrane criteria for systematic review of interventions [31]. This methodology explores the adequacy of sequestration, allocation sequence concealment, blinding of participants and study personnel, blinding for outcome assessment, incomplete outcome or selective outcome reporting, and another potential bias. Any disagreement between the authors was resolved with mutual agreement after discussion.

### Data synthesis and statistical analysis

Outcomes were used in the meta-analysis only if at least three studies reported usable data. The Mantel-Haenszel method for dichotomous data was used to calculate aggregated risk ratios (RRs) with corresponding 95% confidence intervals (CIs). A 2-tailed alpha level of 0.05 was set as the threshold for statistical significance. The I^2^ statistic was used to assess unexplained statistical heterogeneity among studies. A value of between 25% and 50% was considered low heterogeneity, between 50% and 75% moderate heterogeneity, and more than 75% were considered high heterogeneity. The meta-analysis was performed with a fixed effects model, while a random effects model was used if heterogeneity was encountered. We did not examine publication bias because the small number of studies (<10) meant that our meta-analysis was underpowered any such bias. Statistical analysis was performed using Review Manager, version 5.3 (Copenhagen: The Nordic Cochrane Centre, The Cochrane Collaboration, 2014).

## RESULTS

### Search results and characteristics of included trials

Figure 1 shows the PRISMA flow chart summarizing the search strategy. The literature search identified 36 full-text articles, of which 8 RCTs were eligible for inclusion in this study after full read [24-29,32,33]. A total of 6,311 patients were included, of which 3267 patients received tocilizumab and 3,044 received standard care/placebo. Baseline characteristics were similar across the intervention and standard care or placebo groups. Detailed characteristics of the studies are described in **Table 1**.

**Table 1.** Study Characteristics.

| Reference | Groups | Sample size | Median Age (years) | Race or ethnic group – white % | Males (%) | DM2(%) | HTN (%) | CRP (mg/L) | Concomitant treatment |
| --- | --- | --- | --- | --- | --- | --- | --- | --- | --- |
| BACC Bay Tocilizum ab | TCZ | 161 | 61.6 (46.4–69.7) | 44 | 60% | 28 | 50 | 110 (64.9-175.3) | Antivirals<br>Hydroxychloroquine<br>Glucocorticoids |
|  | Placebo | 82 | 56.5 (44.7–67.8) | 40 | 55% | 37 | 46 | 94.3 (58.4-142) |  |
| RCT-TCZ-COVID | TCZ | 60 | 61.5 (51.5-73.5) | N/A | 66.7 | 16.7 | 27 | 105 (50-146) | Hydroxychloroquine<br>Azithromycin<br>Antiretrovirals<br>Anticoagulation |
|  | Standard care | 63 | 60.0 (54-69) | N/A | 56.1 | 13.6 | 29 | 65 (32-118) |  |
| CORIMU NO-TOCI-1 | TCZ | 63 | 64.0 (57.1-74.3) | N/A | 70 | 33 | 33 | 119.5(74.5-219.5) | Antivirals<br>Antiretrovirals<br>Anakinra<br>Eculizumab<br>Glucocorticoids<br>Anticoagulation<br>Azithromycin |
|  | Usual care | 67 | 63.3 (57.1-72.3) | N/A | 66 | 34 | 30 | 127(84-171) |  |
| COVACT A | TCZ | 294 | 60.9 ±14.6 | 59.9 | 69.7 | 35.7 | 60.5 | 168.4±101.4 | Glucocorticoids<br>Antivirals<br>Hydroxychloroquine<br>Convalescent plasma |
|  | Placebo | 144 | 60.6±13.7 | 52.8 | 70.1 | 43.1 | 65.3 | 172.6±114.0 |  |
| REMAP-CAP | TCZ | 353 | 61.5±12.5 | 70 | 74 | N/A | N/A | 150(85-221) | Hydrocortisone<br>Antivirals<br>Immunoglobulin<br>Macrolide<br>Antiplatelet<br>Statins<br>Anticoagulation |
|  | Control | 402 | 61.1±12.8 | 74 | 70 | N/A | N/A | 130(71-208) |  |
| EMPACT A | TCZ | 249 | 56.0±14.3 | 11.2 | 60.2 | N/A | N/A | 124.5 (2.5-2099) | Glucocorticoids<br>Antivirals |
|  | Placebo | 128 | 55.6±14.9 | 15.6 | 57 | N/A | N/A | 143.4(9-3776) |  |
| RECOVERY | TCZ | 2022 | 63.3 (13.7) | 67 | 66 | 28 | 22 | 143(107-204) mg/L | Hydroxychloroquine<br>Antiviral<br>Azithromycin<br>Antiretrovirals |
|  | Usual care | 2094 | 63.9 (13.6) | 68 | 69 | 29 | 24 | 144(106-205) |  |
| TOCIBRAS | TCZ | 65 | 57.4 (15.7) | 68 | N/A | 34 | 46 | 160±104 | Corticosteroids<br>Anticoagulation<br>Other immunosuppressants<br>Hydroxychloroquine<br>Azithromycin |
|  | Control | 64 | 57.5 (13.5) | 69 | N/A | 31 | 53 | 193±283 |  |

**Fig 1.**
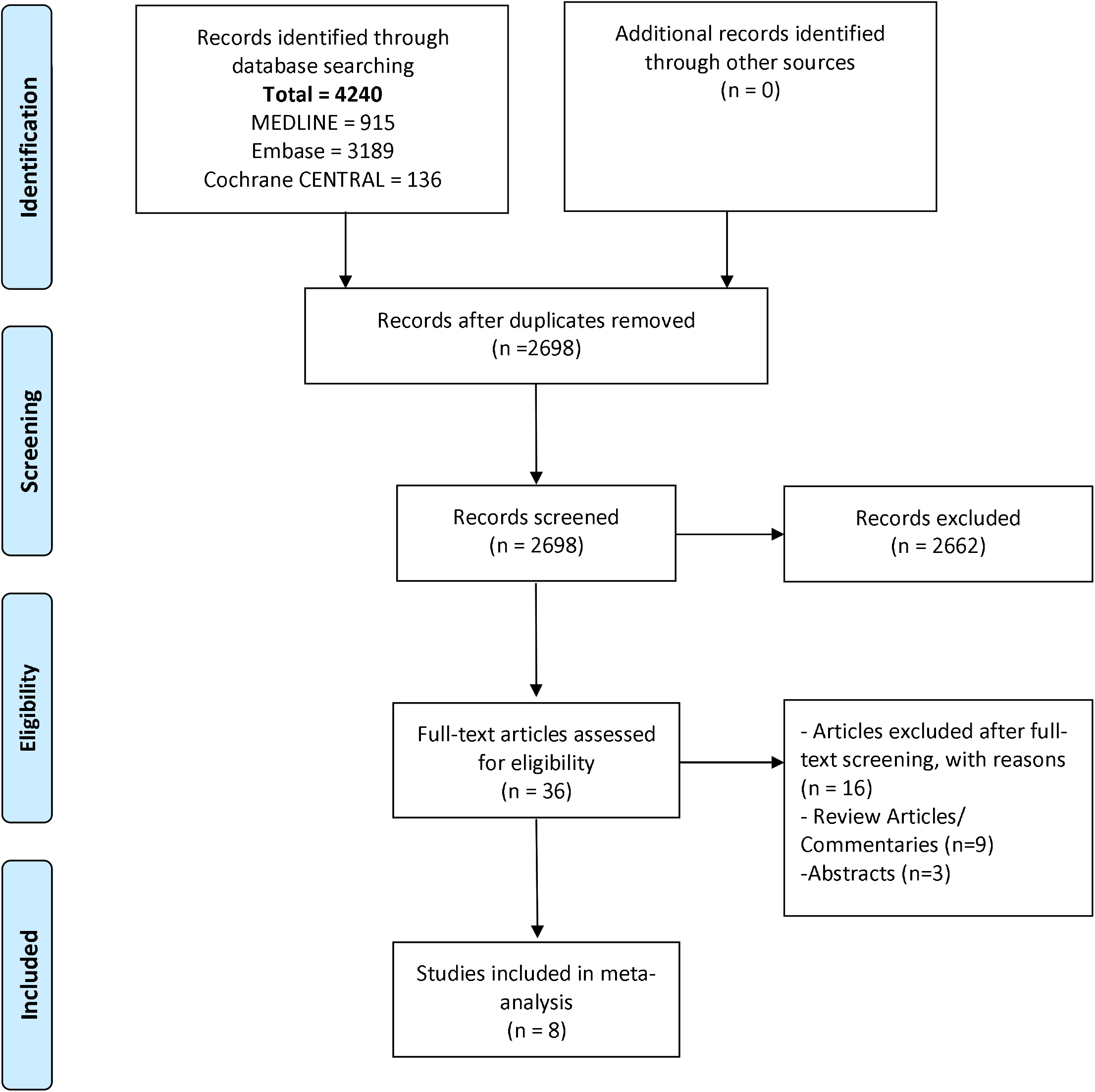
PRISMA flow chart outlining literature search.

All trials included in the analysis studied hospitalized patients with COVID-19 and were multicenter in design. One study was conducted in Italy, one in Brazil, one in France, one in the United Kingdom, and one in the United States of America (USA). Three international trials were conducted across multiple countries in Europe, Mexico, Kenya, South Africa, Peru, and Brazil [25,26,32]. Five trials had open-label design and three were double-blinded. Detailed study designs and study criteria are described in **Table 2**.

**Table 2.** Study Design and Criteria.

| Reference | Site | Design | Dates | Follow up | Inclusion criteria | Primary outcome |
| --- | --- | --- | --- | --- | --- | --- |
| BACC Bay Tocilizumab | USA | Randomized, double-blind, placebo-controlled trial. | April 20 to June 15, 2020 | 28 days | SARS-CoV2+ fever+ pulmonary infiltrates or need for supplemental oxygen. At least one of the following laboratory criteria also had to be fulfilled: a CRP 50 mg/L, ferritin > 500 ng/ml, D-dimer level > 1000 ng/ml, or LDH level > 250 U/L | Intubation (or death, for patients who died before intubation) |
| RCT-TCZ-COVID | Italy | Cohort-embedded, investigator-initiated, multicenter, open-label, randomized clinical trial. | March 31 to June 11, 2020 | 28 days | SARS-CoV2+ fever + (PaO<sub>2</sub>/FiO<sub>2</sub>) ratio between 200 and 300mg Hg, and/or CRP levels of >10 mg/dL and/or CRP level increased to at least twice the admission measurement. | Entry into ICU with invasive mechanical ventilation, death from all causes, or PaO<sub>2</sub>/FiO<sub>2</sub> ratio less than 150 mm Hg |
| CORIMUNO-TOCI-1 | France | Prospective, open label randomized clinical trial. | March 31 to April 18, 2020 | 60 days | SARS-CoV2+ or CT chest findings + moderate, severe, or critical pneumonia O<sub>2</sub> >3 L/min, WHO-CPS score ≥5 | Scores higher than 5 on the WHO-CPS scale on day 4 and survival without need of ventilation (including noninvasive ventilation) at day 14 |
| COVACTA | Global | Multicenter, randomized, double-blind, placebo-controlled phase III trial. | April 3 to May 28, 2020 | 28 days | SARS-CoV2+ chest Xray or CT findings + SpO<sub>2</sub><93% or PaO<sub>2</sub>/FiO<sub>2</sub> ratio<300 | Clinical status at day 28, as assessed on the seven-category ordinal scale. |
| REMAP-CAP | Global | International, adaptive platform, open label randomized controlled trial. | April 19 to November 19,2020 | 90 days | Confirmed COVID-19 who were admitted to the ICU and were receiving respiratory or cardiovascular organ support. | Number of respiratory and cardiovascular organ support-free days up to day 21 |
| EMPACTA | Global | Double-blinded, placebo-controlled, multicenter trial | N/A | 60 days | Confirmed COVID19 along with radiologic features who were saturating below 94% while breathing ambient air. | Mechanical ventilation (invasive mechanical ventilation or extracorporeal membrane oxygenation) or death by day 28 |
| RECOVERY | UK | Randomized, controlled, open-label, platform trial. | N/A | 28 days | Clinical evidence of progressive COVID-19 (defined as oxygen saturation <92% on room air or receiving oxygen therapy, and CRP ≥75 mg/L). | All-cause mortality |
| TOCIBRAS | Brazil | Multicenter, randomized, open label, parallel group, superiority trial. | May 8 to July 17, 2020 | 15 days | Confirmed severe or critical COVID-19 + receiving supplemental or receiving mechanical ventilation for <24 hours before analysis + At least two of the following criteria had to be met: D dimer >1000 ng/mL, CRP >50 mg/L, ferritin >300 µg/L, or LDH greater than the upper limit of normal. | Clinical status at 15 days evaluated with the use of a seven-level ordinal scale |

Importantly, the trials enrolled patients with varying disease severity. The BACC Bay Tocilizumab study excluded patients that required greater than 10L/min oxygen requirement, while the RCT-TCZ-COVID study excluded patients that required non-invasive or invasive mechanical ventilation [24,27]. Similarly, the CORIMUNO-TOCI-1 trial enrolled patients needing >3L/min supplemental oxygen and excluded patients without ventilation or admission to the ICU [28]. The COVACTA, TOCIBRAS, and EMPACTA trials included patients in the ICU and 30%, 32%, and 26.5% of the patients were in the Intensive Care Unit (ICU) or non-ICU with non-invasive ventilation or high flow oxygen [25,26,29]. In the REMAP-CAP study, 42% of the patients were on non-invasive ventilation and 29% on invasive ventilation [32]. In the RECOVERY trial, at the time of second randomization, 562 (14%) patients were receiving invasive mechanical ventilation, and 1686 (41%) patients were receiving non-invasive respiratory support (including high-flow nasal oxygen, continuous positive airway pressure, and non-invasive ventilation) [33].

### Risk of Bias Assessment

The risk of bias assessment for the included trials is presented in **Figure 2**. All trials reported using random sequence generation. Concealment of allocation was not mentioned in five trials and therefore risk of allocation concealment remains unclear in these trials [27-29,32,33]. Also, five trials had open label study design [27-29,32,33]. The risk of performance and selection bias was high in these trials as participants and personnel were not blinded to the assigned treatment. Bias due to selective reporting was deemed high in two studies because they were preliminary reports and data were not presented for some outcomes [32,33]. Risk of attrition bias was deemed high in one trial due to incomplete outcome [29]. In the rest of the studies, risk of attrition bias was low [24-28,32,33].

**Fig 2.**
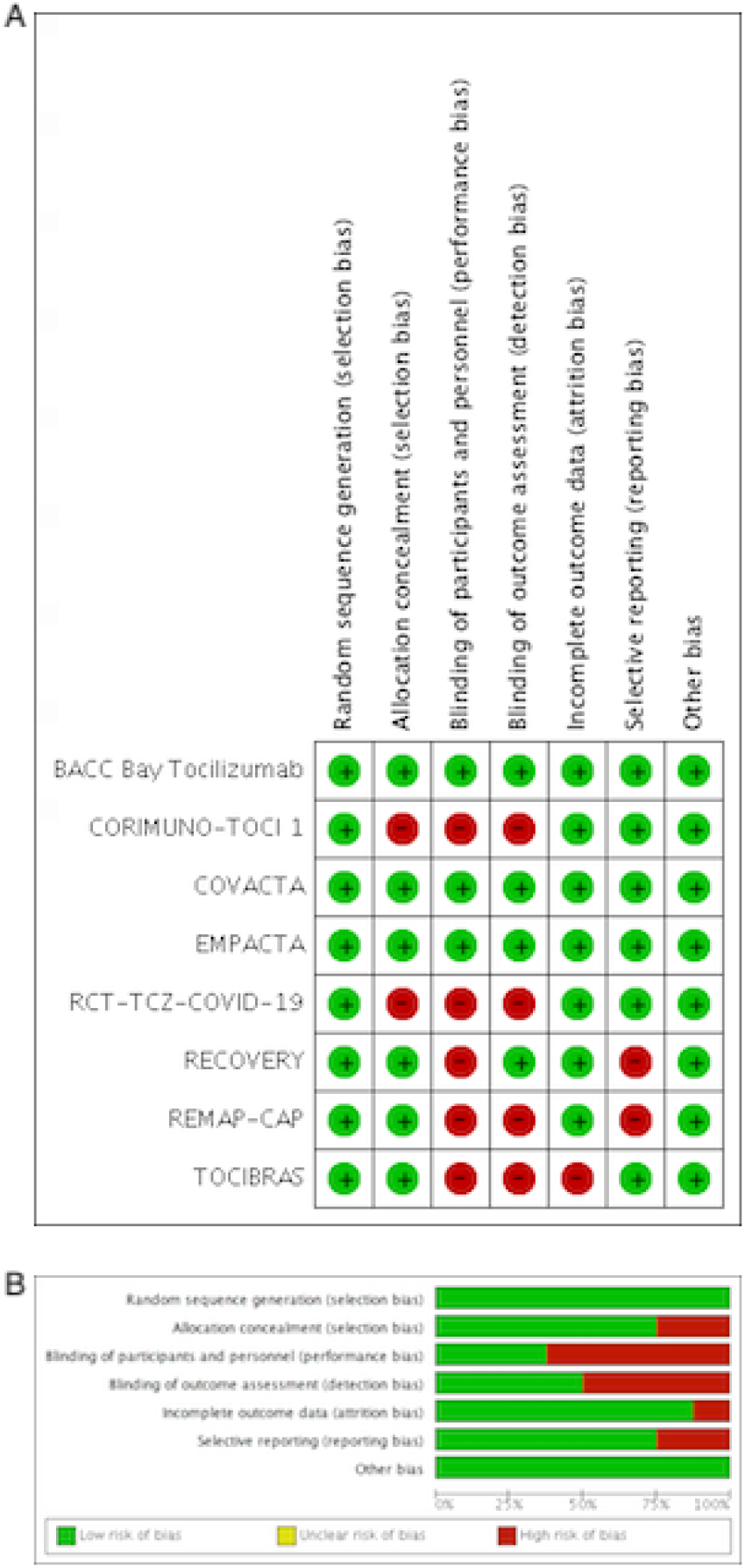
Risk of bias assessment of trials included in the study.

### Assessment of Outcomes

#### All-cause Mortality

All 8 RCTs reported the outcome of all-cause mortality [24-29,32,33]. A total of 810 deaths out of 3267 participants were reported in the tocilizumab arm compared to 893 deaths out of 3044 participants in the standard care/placebo arm. Pooled analysis showed a significant reduction in all-cause mortality at 28 days with tocilizumab therapy than standard therapy or placebo (RR 0.90, 95% CI 0.83-0.97, p=0.009) **(Figure 3A)**.

**Fig 3.**
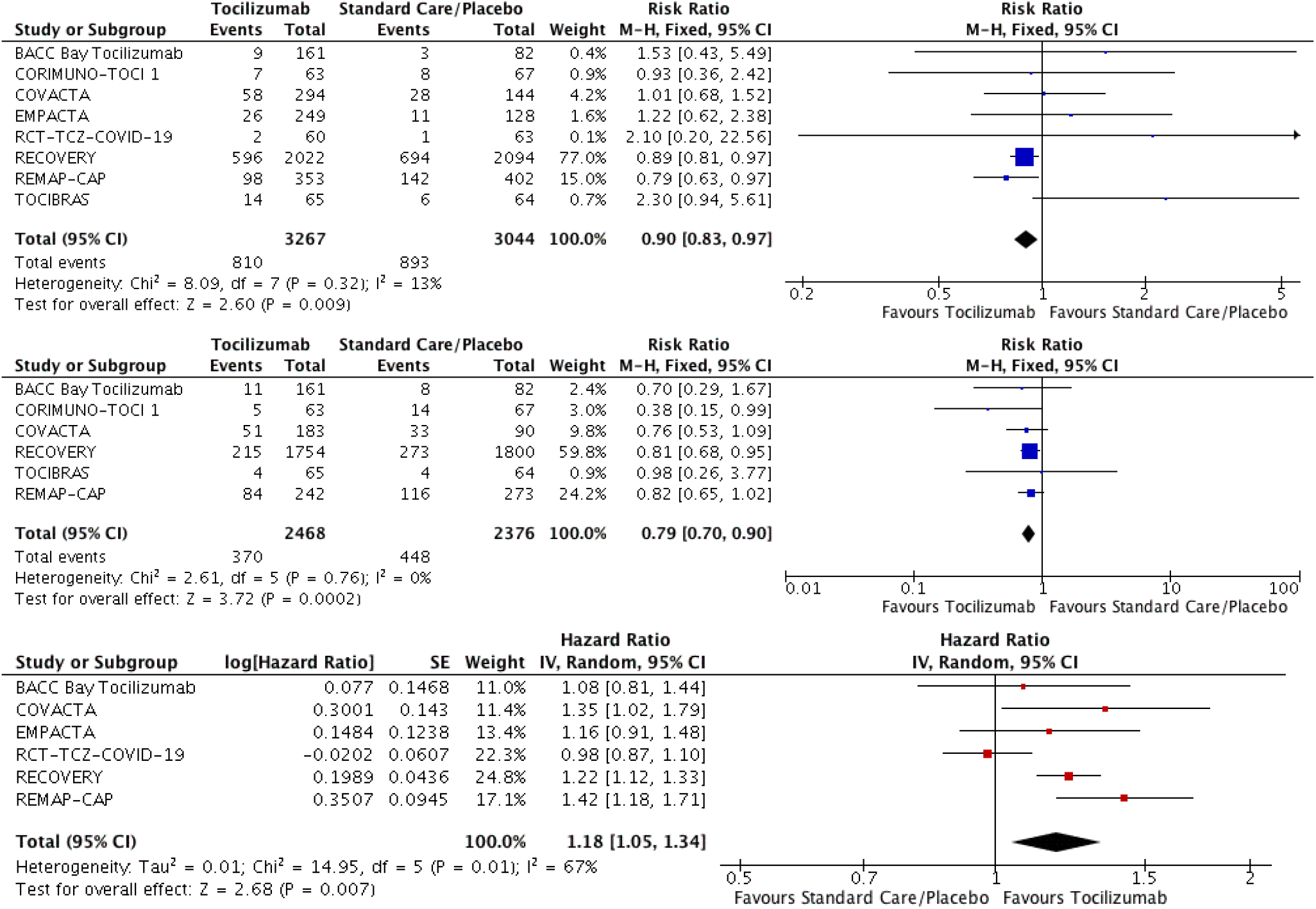
Forest plot for primary and secondary outcomes. A: Mortality outcome B: Progression to mechanical ventilation C: Time to discharge

#### Progression to mechanical ventilation

Six RCTs reported the outcome of need for mechanical ventilation [24,25,28,29,32,33]. 370 out of 2468 participants progressed to mechanical ventilation in the tocilizumab group compared to 448 out of 2376 participants in the other group. Statistical significance was observed with RR of 0.79 and 95% CI 0.70-0.90 with a p value =0.0002 **(Figure 3B)**.

#### Time to Hospital Discharge

Six RCTs reported the outcome of median time to hospital discharge [24-27,32,33]. Pooled analysis showed significantly improved outcome of median time to hospital discharge with tocilizumab therapy than standard therapy or placebo (HR 1.18, 95% CI 1.05-1.34, p=0.007) **(Figure 3C)**.

#### Subgroup analysis of all-cause mortality

##### By the severity of illness

In patients with severe or critical disease (patients on non-invasive or invasive ventilation), a total of 766 deaths out of 2734 participants were reported in the tocilizumab arm compared to 870 deaths out of 2704 participants in the standard care or placebo arm (RR 0.89, 95% CI 0.82-0.96, p=0.005) [25,29,32,33]. In patients with mild or moderate disease, 44 deaths out of 533 participants were reported in the tocilizumab arm compared to 23 deaths out of 340 participants in the standard care or placebo arm (RR 1.21, 95% CI 0.74-1.98, p=0.44) [24,26-28].

##### By sample size

In RCTs with more than 100 patients in each arm, there was a statistically significant reduction in all-cause mortality in patients that received tocilizumab compared to standard care or placebo (RR 0.88, 95% CI 0.82-0.96, p=0.003) [25,26,33,33]. However, in RCTs with less than 100 patients in each arm, all-cause mortality outcomes were not statistically significant (RR 1.56, 95% CI 0.90-2.71, p=0.12) [24,27-29].

##### By trial design

In open-label RCTs, 717 deaths out of 2563 participants were reported in the tocilizumab arm compared to 851 deaths out of 2690 participants in the standard care or placebo arm (RR 0.88, 95% 0.81-0.96, p=0.004) [27-29,32,33]. In double-blinded RCTs, a total of 93 deaths out of 704 participants were reported in the tocilizumab arm compared to 42 deaths out of 324 participants in the other arm (1.10, 0.95% 0.79-1.54, p=0.57) [24-26].

##### By concomitant dexamethasone use

In patients that received dexamethasone along with tocilizumab, a total of 720 deaths out of 2624 participants were reported in the tocilizumab arm compared to 847 deaths out of 2624 participants in the standard care or placebo arm. Pooled analysis showed a significant reduction in all-cause mortality with tocilizumab therapy than standard therapy or placebo (RR 0.88, 95% CI 0.81-0.95, p=0.002) [26,32,33].

##### By the site of study

A pooled analysis showed a trend towards positive outcomes with tocilizumab therapy in global RCTs, although results did not reach statistical significance (RR 0.87, 95% CI 0.72-1.04, p=0.12) [25,26,32]. There was a statistical difference in all-cause mortality outcomes in single-site RCTs in patients that received tocilizumab compared to standard care or placebo (RR 0.91, 95% CI 0.83-0.99, p=0.03) [24,27,28,29,33].

## DISCUSSION

This meta-analysis provides a comprehensive aggregate analysis of the available randomized trials to date on the efficacy and safety of tocilizumab therapy in hospitalized patients with COVID-19 Infection. The results of this study suggest the mortality benefit of tocilizumab treatment in all hospitalized COVID-19 patients and in patients with severe or critical disease. Tocilizumab therapy was also associated with less incidence of progression to mechanical ventilation and possibly with early hospital discharge or readiness to discharge than standard therapy or placebo in hospitalized COVID-19 patients.

Recent Infectious Diseases Society of America (IDSA) guidelines conditionally suggest the use of tocilizumab in addition to standard of care rather than standard care alone among hospitalized patients with progressive severe or critical COVID-19 [34]. Severe illness was defined as patients with SpO2≤’94% on room air, including patients on supplemental oxygen. Critical illness was defined as patients on mechanical ventilation and extracorporeal membrane oxygenation (ECMO). Critical illness also included end organ dysfunction as is seen in sepsis/septic shock.

Our study findings support these guidelines and suggest the efficacy of tocilizumab along with dexamethasone in hospitalized COVID-19 patients. In our meta-analysis, patients who received tocilizumab had reduced need for invasive mechanical ventilation compared to standard care or placebo.

The National Institutes of Health (NIH) guidelines also recommend the use of tocilizumab along with dexamethasone in hospitalized patients that have been admitted to the intensive care unit within the prior 24 hours and who require invasive mechanical ventilation, non-invasive mechanical ventilation, or high-flow nasal cannula oxygen (>0.4 FiO2/30 L/min of oxygen flow). They also recommend use of tocilizumab in hospitalized patients with rapidly increasing oxygen needs and with significantly increased markers of inflammation [35]. Our study findings support these guidelines, especially in patients with severe or critical disease in whom tocilizumab use favored improved mortality, although the results did not reach statistical significance.

The hyperinflammation phase in COVID-19 involves several cytokines and chemokines. However, tocilizumab only inhibits one cytokine, IL-6. In the RCT-TCZ-COVID study, where only 4% of the patient population received steroids. Tocilizumab use did not reduce the risk of clinical worsening in the study population [27]. Also, in the BACC Bay Tocilizumab study, only 10% of the study population received glucocorticoids [24]. There was no significant effect on the risk of intubation or death, on disease worsening, on time to discontinuation of supplemental oxygen. With concomitant dexamethasone administration, there appears to be a synergistic or additive effect on numerous inflammatory pathways. In the EMPACTA trial, 55.4% of the patients in the tocilizumab group and 67.2% of those in the placebo group received concomitant dexamethasone [26]. Patients who received tocilizumab were less likely than those who received placebo to undergo mechanical ventilation or die by day 28 (HR 0.56; 95% confidence interval [CI], 0.33 to 0.97) was no significant difference in overall mortality. In the REMAP-CAP trial, steroid use increased to 88%, and tocilizumab was found to improve mortality and time to clinical improvement [32]. In the RECOVERY trial, 82% of the patients received dexamethasone and tocilizumab resulted in a 6% reduction in mortality when combined with dexamethasone but had no impact on mortality given alone [33].

The severity of illness seems to play an important role in determining the benefit of tocilizumab in this population. In the RCT-TCZ-COVID study, only patients with PaO2/FiO2 ratios between 200 and 300mm were included [27]. In the BACC Bay tocilizumab study, patients requiring >10L/min oxygen were excluded [24]. In both studies, primary outcomes were not met. In the CORIMUNO-TOCI-1 trial, patients with a WHO-CPS score of 5 with O2 levels of 3 L/min or higher but without non-invasive ventilation (NIV) or mechanical ventilation (MV) were enrolled. Survival without invasive or non-invasive mechanical ventilation by day 14 was met, but mortality at day 28 was not different between the groups. Effects of tocilizumab may have also been diminished due to greater steroid use in the control group [28]. In the COVACTA trial, 38% of the patients were mechanically ventilated. There was no significant difference between the tocilizumab and placebo groups concerning clinical status or mortality at day 28, although the time to hospital discharge was shorter with tocilizumab (HR 1.35; 95% CI, 1.02 to 1.79).

Besides, patients initially located outside the ICU were less likely to be transferred to the ICU if treated with tocilizumab [25]. In the REMAP-CAP trial, the greatest benefit was seen in patients admitted to the ICU for organ support (e.g., high-flow nasal cannula or ventilation) [32].

Variations in inflammatory cascade pathophysiology make the timing of initiating treatment with tocilizumab crucial. The RECOVERY trial enrolled patients who have hypoxia with CRP>75 mg/L. There was no difference when comparing patients within <2 days of hospital admission versus >2 days after hospital admission. Treatment with tocilizumab improved survival, probability of discharge from hospital alive by 28 days and reduced the probability of progressing to require invasive mechanical ventilation [33]. In the REMAP-CAP trial, tocilizumab was initiated within 24 hours of organ support in the ICU, although effects were similar across all C-reactive protein subgroups. However, the effects of IL-6 inhibition were strongest and statistically significant among patients with the highest CRP levels [32]. Additional criteria based on clinical trajectory or ICU admission will be necessary to determine the ideal timing for tocilizumab therapy.

One of the limitations of the meta-analysis is integral to the methodology. The summarization of information may ignore the important difference between studies. Secondly, the number of patients in the RECOVERY trial was much higher than other RCTs (study weight 77%) [33]. There were differences in enrollment criteria, the time at which anti–interleukin-6 therapy was initiated, the primary outcome, and background care. Also, five of the included studies had an open label design, implying high risk of performance and selection bias due to lack of blinding of participants and personnel to intervention, limiting our ability to interpret results. Lastly, we included one trial from the preprint databases, which has not been peer-reviewed [33]. Preprint articles possibly indicate the undetermined quality of available literature. One of the trials also ended early due to safety concerns, although mortality in the control arm was extremely low [29].

In conclusion, our meta-analysis suggests tocilizumab, when used along with dexamethasone, could be an effective therapeutic option with promising evidence on reduced mortality, progression to mechanical ventilation, and discharge from hospital. Limitations may exist in terms of obtaining tocilizumab as it is an expensive monoclonal antibody. Supplies may be limited, and many hospitals may not necessarily have adequate quantities. Clinical judgement is also necessary as some patients may not require the use of tocilizumab. Future studies could assess the timing of intervention based on associated co-morbidities and inflammatory markers, the economic benefits of tocilizumab and other IL-6 inhibitors in patient outcomes, and critical healthcare resource usage.

## TAKE HOME MESSAGE

Tocilizumab therapy is associated with reduced mortality, reduced mechanical ventilation progression, and early hospital discharge in hypoxemic COVID-19 patients. Reduced mortality was also observed in patients with severe or critical disease and patients who received concomitant dexamethasone treatment.

## Data Availability

All the data is available within the manuscript

## Declarations

### Funding

None

### Conflicts of interest/Competing interests

On behalf of all authors, the corresponding author states that there is no conflict of interest.

### Availability of data and material

Within manuscript

### Authors’ contributions Author Contributions

VS and MSK led the study and wrote the report. VS, CB, KDA, AF, EM obtained and analyzed data. MSK did the statistical analysis. VS, AL and MSK had full access to raw data. VS, and MSK had full access to, and verified, all data in the study and had final responsibility for the decision to submit for publication.

## Abbreviations

BACC: Boston Area COVID-19 Consortium
RCT-TCZ-COVID-19: Randomized Controlled Trial-Tocilizumab-COVID-19
CORIMUNO-TOCI-1: Cohort Multiple Randomized Controlled Trials Open-label of Immune Modulatory Drugs and Other Treatments in COVID-19 Patients
EMPACTA: Evaluating Minority Patients with Actemra
REMAP-CAP: Randomized, Embedded, Multifactorial Adaptive Platform Trial for Community-Acquired Pneumonia
RECOVERY: Randomised Evaluation of COVID-19 Therapy
MEDLINE: Medical Literature Analysis and Retrieval System Online
EMBASE: Excerpta Medica database

## Notes

### Competing Interest Statement

The authors have declared no competing interest.

### Funding Statement

No funding was obtained

